# Estradiol-based Menopausal Hormone Therapy is associated with improved episodic and prospective memory scores in the Canadian Longitudinal Study of Aging

**DOI:** 10.1101/2025.01.13.25320471

**Authors:** Tanvi A. Puri, Laura Gravelsins, Madeline Wood Alexander, Andrew J. McGovern, Paula Duarte-Guterman, Jennifer S. Rabin, Kelly J. Murphy, Liisa A.M. Galea

## Abstract

**Introduction:** Menopause and menopausal hormone therapy (MHT) can influence cognition in postmenopausal women, but previous literature remains equivocal about their effects. MHT varies based on formulation and route of administration, both of which influence dose of estradiol (E2), the estrogen with the greatest affinity to the estrogen receptor. Transdermal E2 avoids hepatic conversion and results in higher plasma levels of E2 than oral E2 formulations. The cognitive domains of executive functions, episodic memory and prospective memory are diminished with age, but may be differentially sensitive to MHT, dependent on the brain regions recruited during the task. There is a lack of research investigating the effects of the age of menopause and estradiol (E2)-based MHT on different cognitive domains.

**Methods:** Using baseline data from the Canadian Longitudinal Study of Aging, we examined the associations between age of menopause and E2 based MHT on performance in three cognitive domains: episodic memory, prospective memory, and executive functions. Our cohort included 7,251 postmenopausal women who were cognitively healthy, with models adjusted for age, education, and body mass index.

**Results:** Earlier age at menopause was significantly associated with lower scores across all cognitive domains. However, for the executive functions domain, an earlier age of menopause was associated with lower scores only in those with grand parity (4 or more children) and there was a greater effect size among APOE ε4 carriers. We found that transdermal E2 was associated with higher episodic memory scores, whereas oral E2 was associated with higher prospective memory scores compared to no MHT. Neither administration route significantly affected executive function.

**Conclusion:** These results highlight the differential effects of E2-based MHT depending on route of administration and cognitive domain, and underscore the importance of considering age of menopause and individual characteristics such as reproductive history and genotype status. This work provides clarity to inconsistencies in the literature and informs the development of precision medicine approaches for cognitive aging in postmenopausal people.

## Introduction

Menopause marks the onset of reproductive senescence and is associated with reductions of ovarian hormones, particularly estrogens and progesterone in human females (Butler & Santoro, 2011; Harlow et al., 2012). Estrogen and progesterone receptors (ERs and PRs) are located in every organ, including the brain, and menopause is associated with an increase in ER density across certain brain regions (Mosconi et al., 2024). Estrogens and progesterone, play a critical role in regulating brain function, including cognition, and influence neuroplasticity in the hippocampus and frontal cortex (Barha & Galea, 2010; Duarte-Guterman et al., 2015; Frankfurt & Luine, 2015; L. A. M. Galea et al., 2017). Therefore, the menopausal transition is an important period to consider for cognitive and brain changes (Brinton, 2008; Brinton et al., 2015; Nerattini et al., 2023).

The menopausal transition is associated with deficits in episodic memory with mixed findings in working memory and verbal fluency (Maki & Weber, 2021; Weber et al., 2013, 2014), which may be due to the brain region recruited and/or menopause-related characteristics, such as age or type of menopause (e.g., spontaneous vs. surgical) (Hogervorst et al., 2022). Earlier age of menopause is associated with greater cognitive decline, increased risk for dementia, and (Georgakis et al., 2016; W. Hao et al., 2023; Kuh et al., 2018; Liao et al., 2023; Whalley et al., 2004; Wood Alexander et al., 2024). greater Alzheimer’s disease (AD) pathology (Coughlan et al., 2023; Gong et al., 2022) compared to a later age of menopause. Other dementia risk factors may interact with age of menopause to increase AD risk, such as APOEe4, the greatest genetic risk factor for late onset sporadic AD(Meng et al., 2012). Together, these findings suggest that age of menopause can influence female brain health, and should be investigated in the context of evaluating interventions such as menopausal hormone therapy (MHT).

MHT typically consists of exogenous estrogens and progestogens to supplement the menopause-related loss of ovarian hormones to treat menopause-related symptoms (Al-Safi & Santoro, 2014). However, its efficacy for protecting against cognitive symptoms and neurodegenerative disease remains debated (Hogervorst et al., 2000; Kantarci et al., 2018; Maki et al., 2011). MHT comes in varied forms that likely exert different effects on brain and cognition depending on composition of hormones and administration routes. Estradiol (the most potent of the estrogens, E2), and estrone (E1), decline significantly over the course of the menopausal transition, with E1 levels, with E1 exceeding E2 levels post-menopause (Rannevik et al., 2008). E2 dose dependently influences neuroplasticity and certain cognitive domains in both animal models and human females including spatial and non-spatial working memory, and verbal memory (Hamson et al., 2016; Lymer et al., 2024; Maki et al., 2011). However, E1 does not exhibit the same neuroplastic effects as E2, and can inhibit hippocampal neurogenesis and hinder hippocampal-dependent memory depending on dose (Barha & Galea, 2010, 2011; McClure et al., 2013; Yagi et al., 2024). These findings suggest that MHT formulation and dose may influence cognition.

Conjugated equine estrogens (CEE), a form of E1-based MHT, have a lower affinity for ERs (Bhavnani, 2003; Perkins et al., 2017). Meta-analyses suggest CEE have fewer cognitive benefits than E2-based MHTs (Anstead et al., 1997; Hogervorst et al., 2000; Hu & Aizawa, 2003; Ryan et al., 2008). The Kronos Early Estrogen Prevention Study (KEEPS), explored the effects of transdermal E2 versus oral CEE versus Placebo but found no significant effect of either MHT on executive functions or verbal memory. Although no cognitive differences were found there were physiological differences. CEE, but not transdermal E2, increased white matter hyperintensities and ventricular volumes compared to placebo (Kantarci et al., 2018). Indeed, another study found transdermal E2 was associated with fewer white matter hyperintensities compared to placebo (Cote et al., 2023). Thus, both the MHT type and route of administration may be important, a notion further supported by rodent studies showing dose-dependent differential effects of E1 and E2 on cognition and neuroplasticity (Barha & Galea, 2010; Duarte-Guterman et al., 2015; L. A. M. Galea et al., 2018). However, in most animal studies, E2 is administered subcutaneously. Together, these findings suggest that transdermal E2 may have more benefits for hippocampus-based structure and function. Unfortunately, MHT route of administration has largely been overlooked in the literature examining the effects of MHT on cognition.

There is a biological rationale that transdermal E2 will differ in terms of active agent compared to oral E2. Oral E2 is largely converted to E1 via first-pass hepatic metabolism, reducing the bioavailability and efficacy of oral E2 (Gleason et al., 2005; Kopper, 2008; Kuhl, 2005; Lievertz, 1987). Non-oral routes of administration of E2, may offer significant advantages and improved efficacy compared to oral estrogens, as they do not undergo hepatic conversion and present with similar E2:E1 ratios as in premenopausal females (Kuhl, 2005; Lievertz, 1987; Sitruk-Ware et al., 2007). Indeed, studies have found that transdermal E2 does not increase cardiovascular disease risk factors which are often seen with oral E2 (Goldštajn et al., 2022). To our knowledge, no study has compared oral E2 to transdermal E2 on cognitive outcomes. Given their differential metabolism, and eventual potency for ER action, we hypothesize differential effects of E2 MHT based on route of administration. However, to our knowledge no studies have examined the effects of transdermal versus oral E2 on separate cognitive domains postmenopause.

Another important factor is whether MHT may exert varying influences on different cognitive domains and their neural substrates of involvement (M. S. Gazzaniga et al., 2019; Harvey, 2019). For example, executive cognitive processes involving monitoring, alternating attention, and response inhibition are primarily governed by the frontal lobes (Stuss, 2011). Prospective memory processes involving remembering to carry out future intentions at a particular time or event require involvement of the frontal and medial-temporal lobes, respectively (Simard et al., 2019, McFarland & Glisky, 2009). Episodic memory involved in the acquisition and retention of new information is governed by the medial temporal lobes, particularly the hippocampus (Moscovitch et al., 2016; Nyberg, 2017). ERs are located throughout the medial temporal lobes, and especially the hippocampus, and the frontal lobes, and there is increased ER density in the frontal cortex, but not hippocampus, after menopause or ovariectomy in both humans and rodents (González et al., 2007; Khayum et al., 2014; Mosconi et al., 2024; Perlman et al., 2005). Furthermore, subcutaneous E2 regulates hippocampus structure and function in rodent models (Barha & Galea, 2010; Cardona-Gomez et al., 2004; Gould et al., 1990; Lee, Eid, et al., 2024; Puri et al., 2024; Sheppard et al., 2022; Tuscher et al., 2016; Yagi et al., 2017). MHT with varying levels of estrogens, and routes of administration may differentially affect cognitive processes that depend on brain regions with ER densities susceptible to the influence of menopause. Therefore, The objectives of the present study were to describe how MHT route of administration and age of menopause onset affects cognitive performance across episodic memory, executive functions, and prospective memory.

We hypothesized that an earlier age of menopause onset would be associated with poorer cognitive scores that may differ depending on the cognitive domain. Because of the known differences in cognition with age of menopause (Wood Alexander et al., 2024), we controlled for age of menopause in further analyses on MHT use. We focussed on E2-based forms of MHT, and hypothesized that the route of administration would affect cognitive performance depending on cognitive domain, with more pronounced effects observed in the episodic memory domain, given findings in animal models (Galea et al., 2017) and the plethora of ER in the neural substrates supporting memory function. We also anticipated that transdermal E2 would provide greater cognitive benefits compared to oral E2, with these effects varying by cognitive domain due to postmenopausal changes in ER density in the frontal cortex (Mosconi et al., 2024). Given that both APOE genotype and parity have been linked to the effects of E2 and to AD risk (Barha & Galea, 2011; Corbo et al., 2007; Coughlan et al., 2023; Puri et al., 2024) we also examined the moderating effects of these two factors in our analyses.

## Methods

### CLSA Participants

We used data from the comprehensive cohort of the Canadian Longitudinal Study on Aging (CLSA), a population-based observational study that collected data from men and women aged 45 to 85 years in Canada (Raina et al., 2019; Raina et al., 2009; Tuokko et al., 2017) details of the CLSA’s participant profile and study design have been reported elsewhere (Raina et al., 2019; Raina et al., 2009). Participation in the study was voluntary, and participants were randomly recruited from areas within a 25-50 km radius of eleven collection sites across Canada (Victoria, Vancouver, Surrey, Calgary, Winnipeg, Hamilton, Ottawa, Montreal, Sherbrooke, Halifax, and St. John’s). Individuals were recruited using provincial healthcare registration databases, or via random telephone digit dialing. Of those contacted and eligible, 10% responded, and approximately 45% of those respondents agreed to participate in data collection (Raina et al., 2019). Genomic data was extracted from the CLSA database specifically for APOE genotype (Forgetta et al., 2022).

The data reported in our study are from the baseline survey, in which 30,097 participants completed questionnaires, underwent cognitive testing, provided biological specimens, and underwent detailed physical assessments between 2012 and 2015. The CLSA has ethics approval in all provinces and centers where data collection took place, and all participants were required to provide written informed consent. Our study was also approved by the Research Ethics Board at the University of British Columbia (H20-02663).

### Inclusion and exclusion criteria

The CLSA applies specific inclusion and exclusion criteria, and these are detailed in Raina et al., 2019. Key exclusions include residents of the Canadian Territories and in remote regions, individuals living on First Nations reserves and settlements, and individuals with cognitive impairment at baseline. The CLSA defines cognitive impairment as the inability to independently complete interviews or hear and respond to questions, without further classification beyond this criterion. Protocols are available online from CLSA (https://www.clsa-elcv.ca/).

All participants self-reported their sex at birth as “male”, “female”, or “do not know”. For these analyses on menopause and menopausal hormonal therapies, we included only individuals who reported their sex at birth as female and who self-identified as female. Of all 15,313 females available for inclusion, those that had N/A values for all cognitive tests were excluded. Participants who reported being formally diagnosed with depression or AD were excluded (Depression: n = 3,210, 21%; AD n = 34, 0.22%), as previous work shows individuals with depression and AD show impairments in hippocampal integrity (Campbell & Macqueen, 2004; Chen et al., 2019). Furthermore, only individuals that completed the cognitive tests in English were included (excluded n = 1,165, 7.6%) as test language can influence scores on language-based tests (Tuokko et al., 2017).

At baseline, female participants reported whether they were menopausal or not, defined as “menstrual periods stopped for at least one year and did not restart”. Individuals that identified as menopausal and reported an age of menopause onset were included. Those with a hysterectomy were not excluded. In line with previous work (Wood Alexander et al., 2024) we excluded those who reported an unlikely age of spontaneous menopause (<35 or >65 years, n = 344, 2.24%). Participant exclusions are summarized in Table 1.

**Table 1:** List of cognitive tests and measures used for the executive function, prospective memory, and episodic memory composites.

| Composite | Executive Functions Composite | Prospective Memory Composite | Episodic Memory Composite |
| --- | --- | --- | --- |
| Cognitive Assessments and associated dependent variables | **Choice Reaction Time Test**<br>• Mean reaction time<br>• Mean reaction time to correct response<br>**Stroop Task**<br>• Latency<br>• Number of Errors | **Prospective Memory Task**<br>• Event based prospective memory task<br>• Time based prospective<br>memory task | **RAVLT**<br>• 1-min immediate recall<br>• 5-min delayed recall |

### Menopausal Hormone Therapy

The CLSA provides a list of hormone therapy options for participants to self-select. Participants selected whether they took “estrogen (e.g., premarin and estrace)”, “progesterone (e.g., prometrium and provera)”, “both estrogen and progesterone”, “estrogen gel or cream applied to the skin”, or “intrauterine device with progesterone”. To obtain information on the *type* of estrogen that was being taken (i.e., E2, estrone (E1)) at the time of cognitive testing, we used the exhaustive list of medications. Participants reported all their medications with interviewers, who then linked medications with a corresponding Canadian Drug Identification Number. We classified individuals as currently taking E2 (only E2, or E2 and progesterone/progestin). Participants were then categorized into the following groups based on <u>current</u> MHT use: oral E2 based MHT, transdermal E2 based MHT (including both creams and vaginal MHTs), and never taken any form of MHT. All current E2-based MHT users, regardless of progestogens use, were included.

### APOE e4 and Number of Children Interaction Terms

Carrying one or two copies of the APOE e4 allele increases risk for AD severalfold, and is determined by the single nucleotide polymorphism (SNP) rs429358 (Rubinsztein & Easton, 1999). The polymorphism defined as the APOE e4 allele lies in the fourth exon of the APOE allele, and affects the amino acid at position 130. Carriers of the e4 allele were defined as those that carried the ‘C’ amino acid on SNP rs429358 at least one chromosome.

Our analyses consider interactions of number of children with cognition. Grand parity, i.e., individuals with four or more children, show increased risk for AD, and greater cognitive decline in middle age. Therefore, in line with previous work, we binned individuals with 0, 1, 2, 3, and 4+ children to investigate the effects of grand parity (Bae et al., 2020; Jung et al., 2020; Zhang et al., 2023).

### Cognitive Assessments

The CLSA assessed cognition through a neuropsychological battery of tests that was conducted at baseline. We analysed participants performance on a modified version of the Rey Auditory Verbal Learning Test (RAVLT, 1 minute immediate recall and 5 minute delayed recall (Rey, 1964), Choice Reaction Time Test (Gallacher et al., 2013), the Stroop Test (Bayard et al., 2011; Troyer et al., 2006), the Event-based Prospective Memory Test (PMT), and Timed Prospective Memory Test (TMT) (Loewenstein & Acevedo, 2004). Details on how the cognitive tests were administered have been described elsewhere (Tuokko et al., 2017). These tests were chosen because they sample a variety of cognitive processes dependent on neural substrates potentially sensitive to change in ER density, and because the measures do not have floor and ceiling effects when tested in persons between ages 45 and 85 (Tuokko et al., 2017). The event- and time-based prospective memory tests both evaluate types of prospective memory and rely on the integrity of the frontal lobe, the parietal lobe and the hippocampus (Burgess et al., 2001, 2011; Martin et al., 2007). The Stroop and choice reaction time tests are associated with executive functions (Simard et al., 2019; Tuokko et al., 2017). The RAVLT provides an assessment of episodic memory.

To determine if the tests chosen could be grouped into composite scores we performed a principal component analysis (PCA) on 8 standardized cognitive measures, including performance metrics. Prior to analysis, the data was scaled, centered, and rows containing missing values were removed. To identify clusters of related cognitive measures, hierarchical clustering was performed on the cognitive test contribution loadings of the first two principal components using Ward’s method with Euclidean distance. The optimal number of clusters was determined through silhouette analysis, evaluating solutions ranging from 2 to 7 clusters. The final clustering solution was visualized using a dendrogram with color-coded branches representing clustered regions. Based on this analysis, we created three cognitive composite scores - an episodic memory composite, a prospective memory composite, and an executive functions composite - to investigate different cognitive domains (Table 1). Z-scores were computed using the analytic sample means and standard deviation. These z scores were formatted such that better performance is represented by a higher score. Thus, tests where a numerically higher score is worse were reverse scored. The z-scores of the relevant tests for each cognitive composite were averaged.

Each of these memory domains identified above rely on the integrity of different brain regions. Episodic memory was assessed using the RAVLT 1 minute immediate recall recall and 5-minute delayed recall, and is primarily reliant on medial temporal lobe integrity, especially the hippocampus (Fernaeus et al., 2013). The episodic memory domain may be particularly sensitive to E2, given work in animal models and in humans, (Gervais et al., 2022; Lee, Eid, et al., 2024; Perović et al., 2023). We incorporated executive functions that were measured via the choice reaction time test and Stroop tasks. Executive functions are governed by the prefrontal cortex, as demonstrated in functional neuroimaging studies and lesion studies (Laird et al., 2005; Vendrell et al., 1995). An executive functions composite was included because ER density in the inferior and middle frontal cortex is increased postmenopause (Mosconi et al., 2024) and because many studies on MHT effects on cognition have examined executive functions (Fisher et al., 2018; Gleason et al., 2005; Kantarci et al., 2018). Prospective memory was evaluated using event-based and time-based tests, and is governed both by the frontal and medial temporal lobes (Burgess et al., 2001; Koo et al., 2022; Nurdal et al., 2020). Prospective memory was of interest as there is a postmenopausal increase in ER density in brain regions like the caudate nucleus, which is associated with planning and memory for future tasks (Barkhoudarian & Kelly, 2017; Grahn et al., 2008). This approach allows us to examine a wide spectrum of cognitive processes that could be influenced by hormonal changes in middle age. By examining these different composites, we provide an assessment of diverse cognitive domains which allow us to capture any potential differences in how menopause-related factors and MHT affect different facets of cognition that recruit different brain regions.

### Statistical Analysis

All analyses were performed in R (version 4.3.3; last updated 2024/02/29). Means (and standard deviation(SD)) and frequency statistics were used to summarize the distribution of continuous and categorical variables in the data (ANOVA and ⍰^2^ respectively). Means (SD) and frequencies are reported in Table 2. Linear regression models were used to test associations between cognitive performance and the age of menopause onset as well as the type of E2 based hormone therapy at baseline data collection. Analyses for both age of menopause and E2 MHT were also run with APOE carrier status and previous parity as interaction terms. All models used in these analyses were adjusted for age at testing, years of education, and body mass index (BMI). Analyses on the effect of E2 based hormone therapy also adjusted for the age of menopause. All *p* values were 2-sided and significance threshold was set at *p < 0.05.* All post-hoc analyses accounted for multiple comparisons by using the Benjamini-Hochberg procedure to account for the false discovery rate.

**Table 2:**
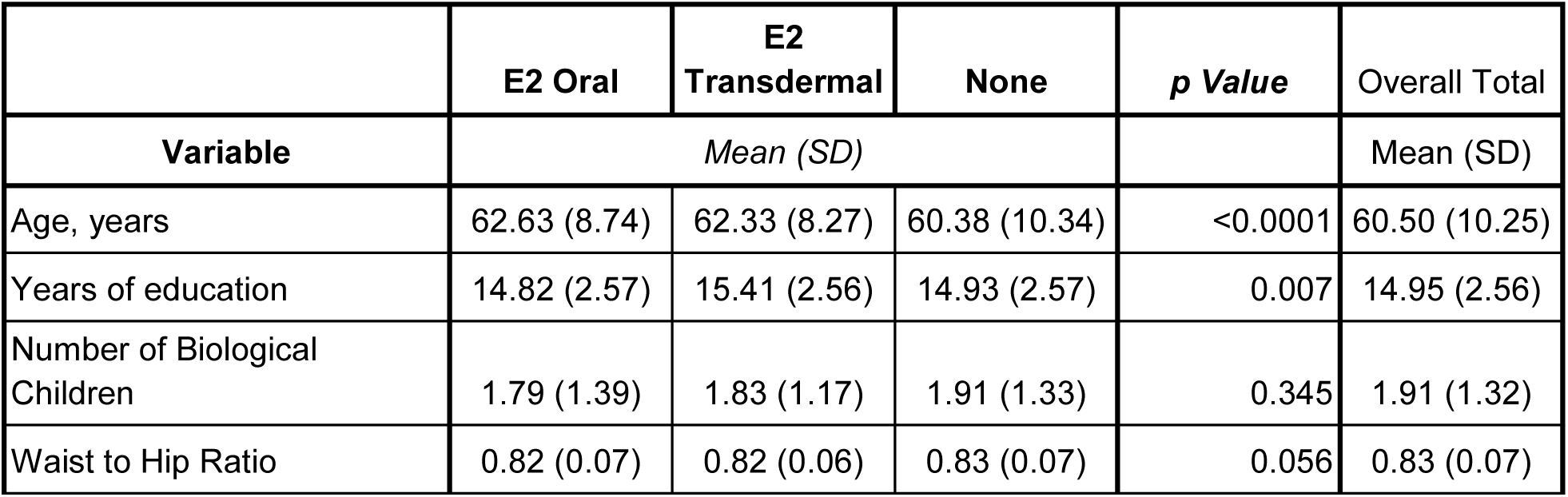
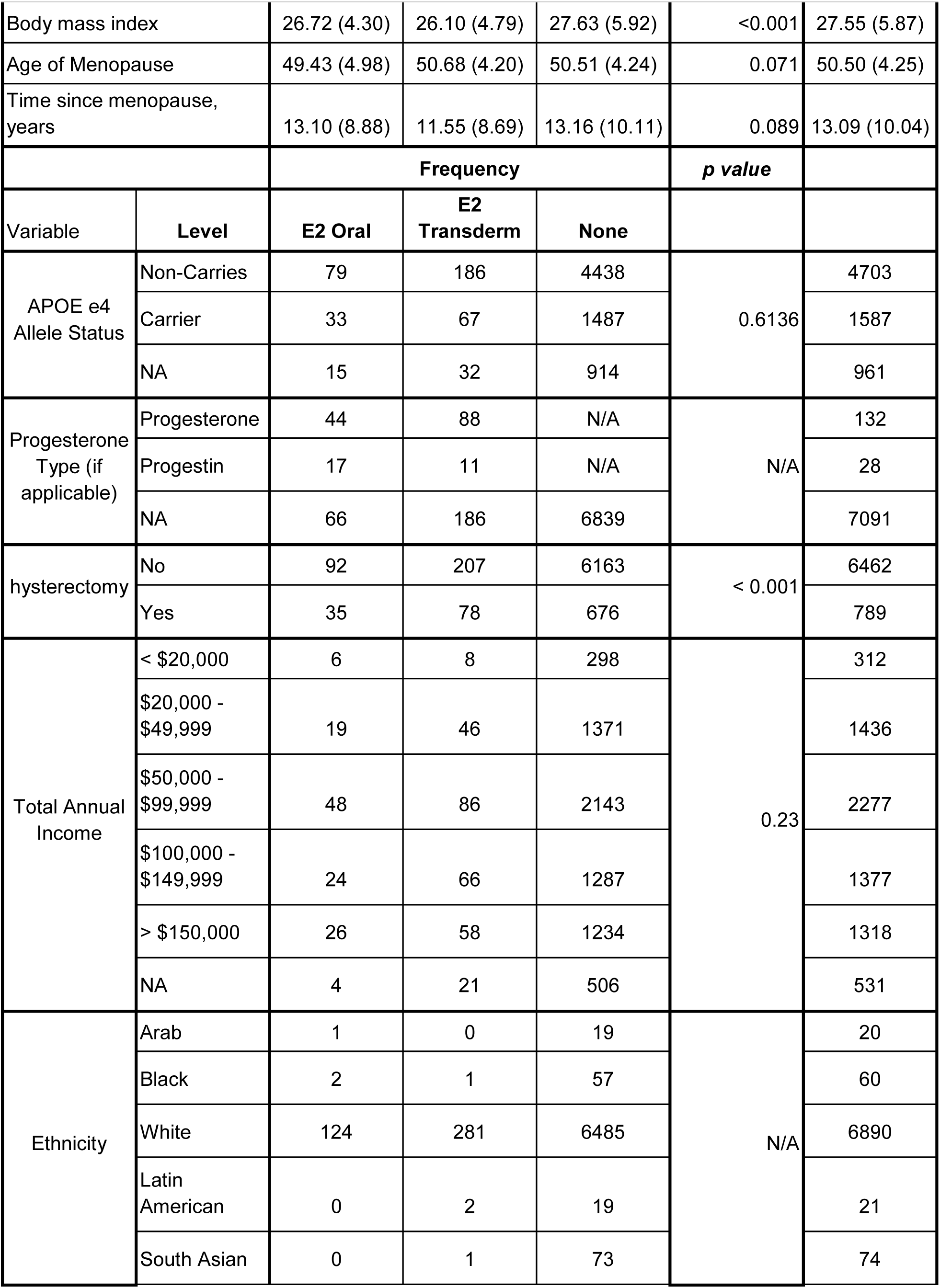
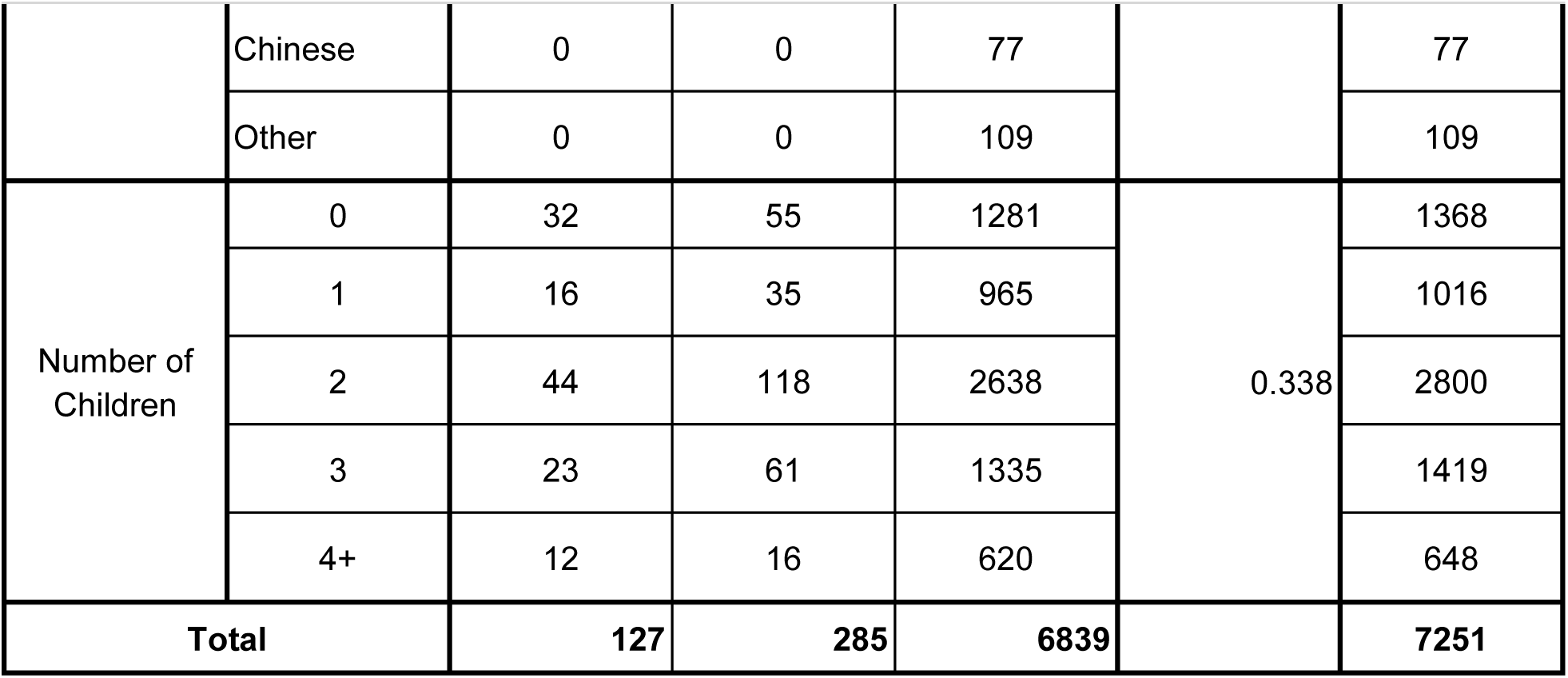
Demographics for postmenopausal women sorted by who have never taken MHT, or are taking oral or transdermal E2. * = ‘Other’ includes individuals who reported their ethnicity as: Chinese (n = 77), Filipino (n = 18), Japanese (n = 11), Korean (n = 2), Southeast Asian (n = 16), West Asian (n = 10), Algonquin Group (n = 1), Canadian (n = 7), Caribbean (n = 1), East Asian (n = 1), Euranian (n = 1), Eurasian (n = 2), Italian (n = 1), Jamaican (n = 2), Judaic (n = 1), Mixed/Mixed Race (n = 5), Native (n = 3), West Indies/Trinidadian (4), Don’t Know/NA (n = 23).

### Data Availability

All data are available through the CLSA (clsa-elcv.ca) to any researchers who meet the criteria for access to the de-identified data.

## Results

### Participant Characteristics

Table 1 summarizes participant demographic characteristics based on whether they were receiving E2 based MHT and route of administration of MHT. Of the 10,291 females, who adhered to all the criteria, our analysis included 7251 individuals who reported using transdermal E2 or oral E2 at the time of testing, or those who reported never using MHT. Significant differences were observed in age, years of education, and body mass index across individuals who took transdermal E2, oral E2, or never took MHT. Age differed significantly across groups (F(2,7248) = 7.75; p < 0.001; partial ⍰^2^ = 0.002), with participants who had never used MHT being younger on average than those in the oral E2 (p = 0.022; Cohen’s *d* = −0.218) and transdermal E2 (p = 0.005; Cohen’s *d* = −0.190) groups. Years of education also differed significantly between groups (F(2,7246) = 4.91; p < 0.007; partial ⍰^2^ = 0.001), with the transdermal E2 group reporting slightly higher levels of education compared to the oral E2 (p = 0.047; Cohen’s *d* = 0.231) and non-MHT (p = 0.006; Cohen’s *d* = 0.186) groups. BMI was significantly different between groups (F(2,7211) = 2.64; p < 0.001; partial ⍰^2^ = 0.003), with the non-MHT group exhibiting a higher BMI compared to the oral E2 (p = 0.127; Cohen’s *d* = 0.154) and transdermal E2 (p < 0.001; Cohen’s *d* = 0.261) groups. Given these significant differences, we adjusted for age, education, and BMI in subsequent analyses.

#### An earlier age of menopause is related to lower performance scores on all memory composites tested–episodic memory composite, prospective memory composite and executive functions composite

##### Episodic Memory Composite

Across all participants, an earlier age of menopause was associated with reduced performance on the episodic memory composite score (F(4,6448) = 302.2; p < 0.001;standardized ⍰ = 0.049)[Figure 2A and D]. We also examined whether being an APOE ε4 carrier or parity (number of biological children) influenced the effect of age of menopause on cognitive outcomes. Neither carrying the APOE ε4 allele nor the number of children interacted with age at menopause to affect performance on the medial temporal lobe composite score (p = 0.960 and 0.929 respectively).

**Figure 1:**
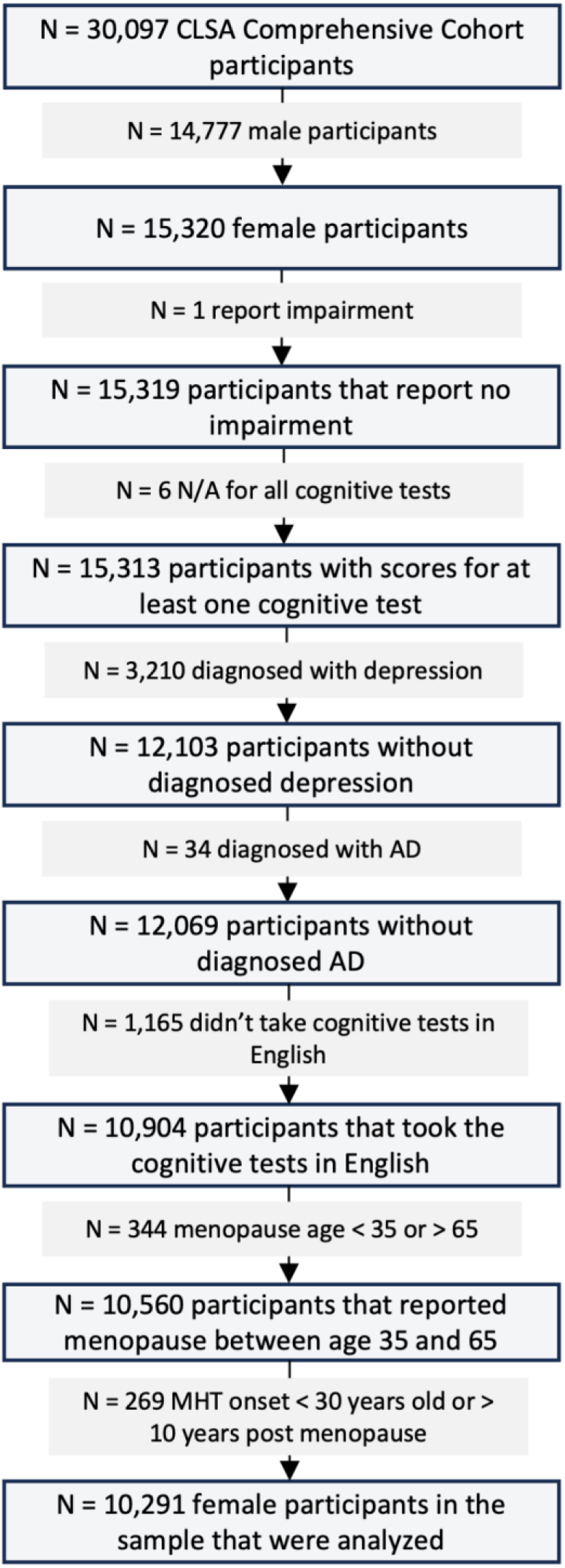
Participant Inclusion Criteria. Participant flow chart depicting our sample selection for analysis of the effects of age of menopause, and E2-based MHT.

**Figure 2:**
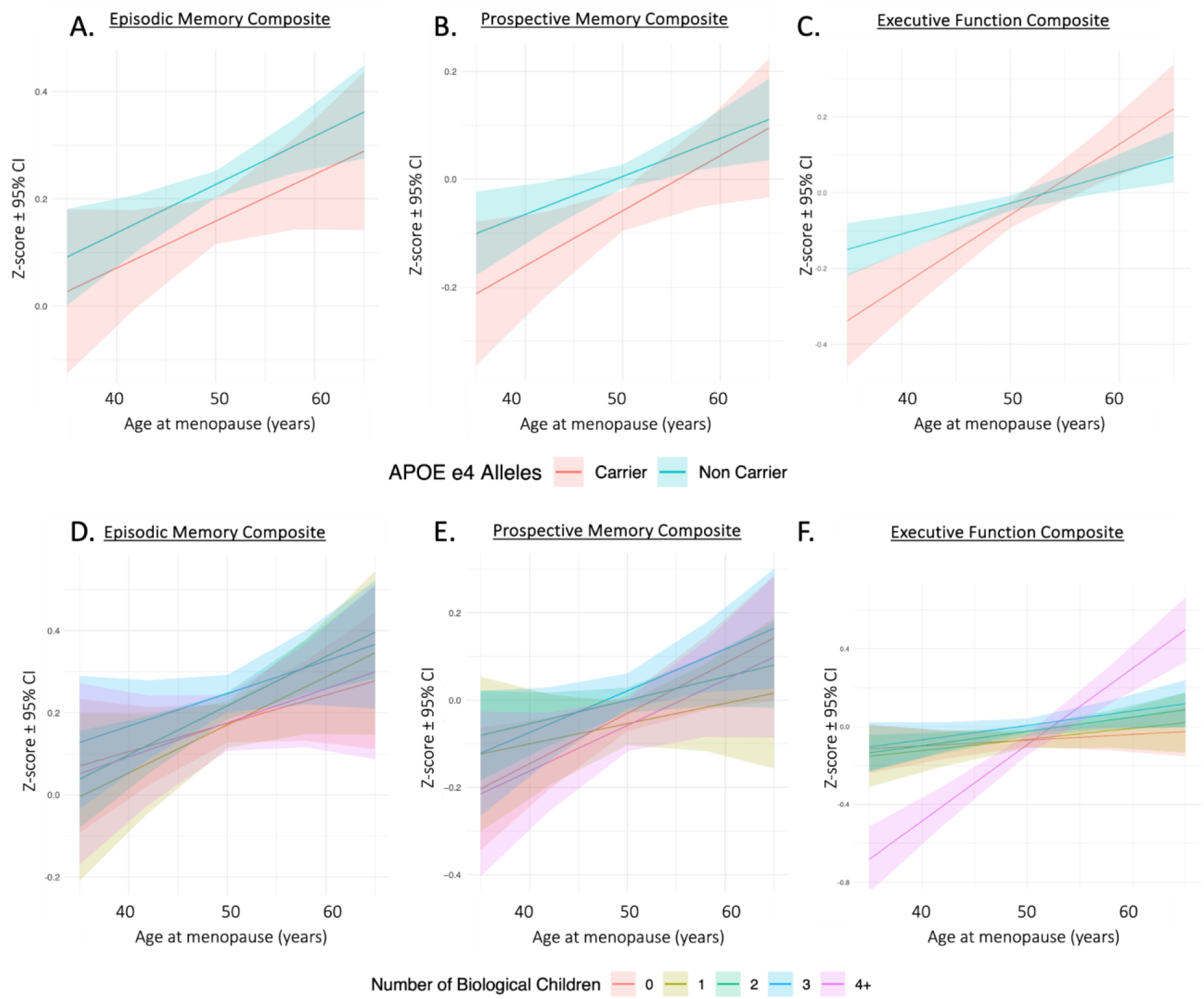
(A-F) Z-score ± 95% CI of cognitive performance on the episodic memory composite score (A,D), prospective memory composite score (B, E), and executive functions composite score (C, F) by APOE ε4 (A-C), and number of children (D-F). (‘C) In APOE ε4 carriers, executive functions based composite scores are associated with worse performance with an earlier age of menopause than non-carriers. (F) In individuals with 4 or more children, executive functions composite scores are associated with worse performance with an earlier age of menopause than those with 0-3 children.

##### Prospective Memory Composite

An earlier age of menopause was also associated with reduced performance on the prospective memory composite score (F(4,6551 = 152.9; p < 0.001; standardized ⍰ = 0.048)[Figure 2B and E]. Similar to the medial temporal lobe composite, neither carrying one or two copies of the APOE ε4 allele nor the number of children interacted with menopause age to affect the prospective memory composite score (p = 0.519 and 0.764 respectively).

##### Executive Functions composite

A slightly different pattern with respect to menopause age was seen on the executive functions composite scores, as an earlier age of memory was associated with reduced scores (F(4,6603) = 380.0; p = <0.001; standardized ⍰ = 0.063)[Figure 2 C and F] but this was significant only in those with four or more children and with a stronger effect size in APOE ε4 carriers.

Carrying one or two copies of the APOE ε4 allele interacted with the age of menopause to affect performance on the executive functions composite score (F(6,5804) = 289.4; p < 0.001)[Figure 2C], such that individuals carrying one or two copies of the APOE ε4 allele had lower scores on the executive functions composite with an earlier age of menopause (p = 0.020; Carrier standardized ⍰ = 0.646) compared to non-carriers (Non-Carrier standardized ⍰ = 0.329) The age of menopause interacted with the number of children an individual had to affect performance on the executive functions composite score (F(12, 6594) = 131.6; p < 0.001)[Figure 2F]. Those with four or more children performed better on the executive functions composite with an older age of menopause, (p < 0.001; standardized ⍰ = 0.733), but those with fewer than four biological children or no children did not (all p’s > 0.442; standardized ⍰ = 0.065 - 0.149).

#### E2-based MHT, depending on route of administration, is associated with improved performance on the episodic and prospective memory composites, but not on the executive functions composite measures

Due to the significant effects on the age of menopause on all memory domains tested, all subsequent analyses used age of menopause as a covariate, along with age, education and BMI.

##### Episodic Memory Composite

Accounting for age of menopause, the type of E2-based MHT affected cognitive performance on the episodic memory composite (F(6,4446) = 149.8, p <0.001; partial ⍰^2^ = 0.005) [Figure 3A]. Those taking Transdermal E2 at the time of the cognitive testing showed improved performance compared to those who have never taken MHTs (p = 0.004; E2 Transdermal CI: 0.302 - 0.541; No MHT CI: 0.197 - 0.247; Cohen’s d = 4.516). In contrast, individuals taking Oral E2-based MHTs performed no differently than those who have never taken MHT (p = 0.395; E2 Oral CI: 0.115 - 0.496; Oral E2 - No MHT Cohen’s d = 1.202).

**Figure 3:**
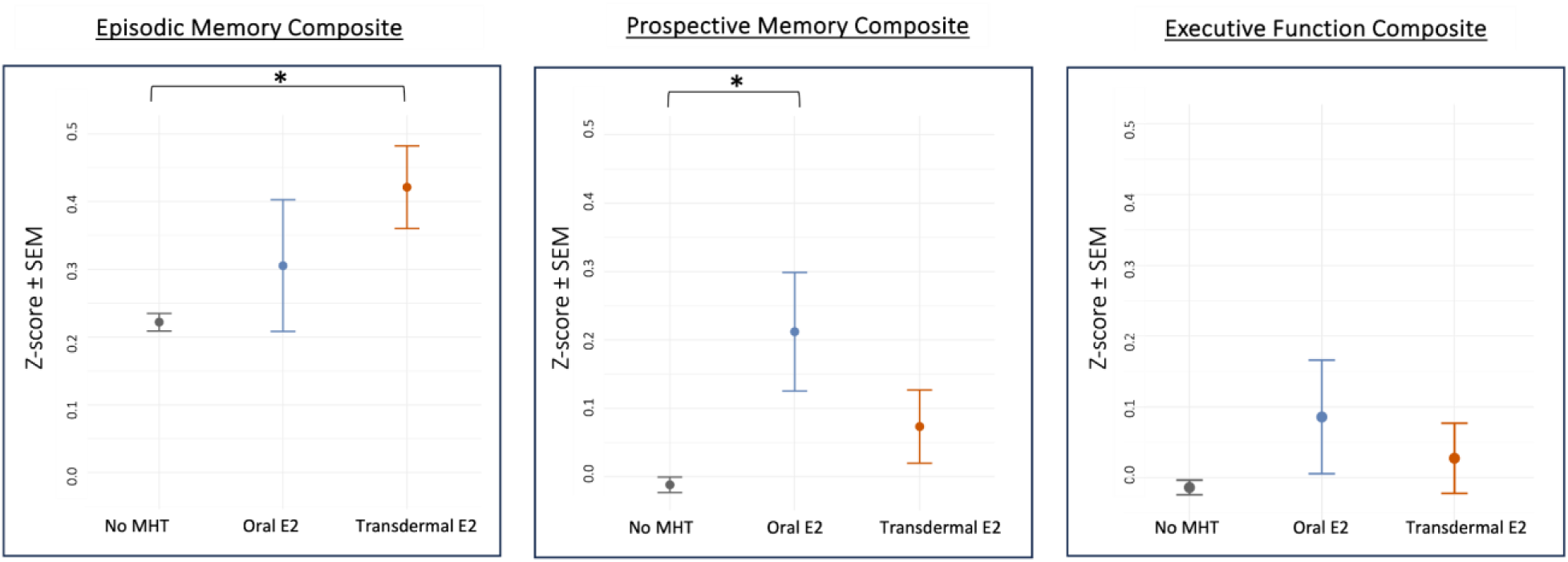
(A-C) Z-score ± SEM of how MHT affects cognitive performance on the episodic memory composite score (A), prospective memory composite score (B), and executive functions composite score (C). (A) Transdermal E2 is associated with improved performance on the episodic memory composite score. (B) Oral E2 is associated with improved performance on the prospective memory composite score. Asterisk indicates p < 0.05.

##### Prospective Memory Composite

Type of E2-MHT also affected prospective memory based cognition accounting for age of menopause (F(6,4557) = 89.32; p = 0.012; partial ⍰^2^ = 0.002)[Figure 3B]. Individuals taking oral E2 performed better than those who have never taken MHTs (E2 Oral-No MHT: p = 0.031; E2 Oral CI: 0.042 - 0.382; No MHT CI: 0.034 - 0.010; Cohen’s d = 3.621). However, those taking transdermal E2 did not significantly differ from those who have never taken MHTs (E2 Transdermal-No MHT: p = 0.173; E2 Transdermal CI: 0.032 - 0.178; Transdermal E2-No MHT Cohen’s d = 2.200).

##### Executive Functions composite

For the executive functions composite score, E2-based MHT, regardless of route of administration, had no significant effect on the composite score (F(6,4553) = 182.5; p = 0.345; partial ⍰^2^ = 0.001; E2 Oral CI: −0.072 - 0.243; E2 Transdermal CI: −0.070 - 0.125; No MHT CI: −0.034 - 0.007; E2 Oral-No MHT (p = 0.536; Cohen’s d = 1.741); Transdermal E2-No MHT (p = 0.536; Cohen’s d = 1.152)[Figure 3C].

##### Neither carrying the APOE ε4 allele nor the number of children modified the effects of E2-based MHT in any memory domain tested

Carrying the APOE ε4 allele (either one or two copies) did not modify the effect of E2-based MHT on the any of the memory composite scores (episodic memory composite: *p = 0.753*; prospective memory composite: *p = 0.605*; executive functions composite: *p = 0.941*). The number of children an individual had did not modify the effect of E2-based MHT on any of the memory composite scores (episodic memory composite: *p = 0.544*; prospective memory composite: *p = 0.870*; executive functions composite: *p = 0.939*). [Supplemental Figure S1]

##### Hysterectomy and taking progestogens did not alter the effects of E2-based MHT

The proportion of females with hysterectomy was similar between the transdermal E2 and oral E2 groups but significantly higher in both MHT groups compared to the group not taking any MHT. In order to account for any confounding of our results, we performed a sensitivity analysis by excluding individuals with a history of hysterectomy, and our findings remained robust and unchanged. We also conducted a sensitivity analysis to assess the impact of progestogens when used in conjunction with E2. After accounting for this variable, our findings remain consistent, demonstrating that the associations between MHT and cognitive domain outcomes were not significantly altered by the inclusion or exclusion of participants using different progestogens.

## Discussion

Our findings show that E2 based MHT are positively associated with prospective and episodic-based memory in postmenopausal individuals. Transdermal E2, but not oral E2, was significantly associated with better episodic memory performance, whereas oral, but not transdermal E2 was significantly associated with better prospective memory performance scores. E2 (regardless of route of administration) had no significant effect on the composite score representing executive functioning. In each of these analyses we corrected for age of menopause, and our findings that E2 can positively affect certain memory domains dependent on route of administration may shed light on inconsistencies in the literature surrounding the equivocal effects of MHT on cognition. In this cohort of postmenopausal adult females in Canada, we found that an earlier age of menopause reduced cognitive performance on all cognitive domains tested, which is largely consistent with past literature. However, age of menopause did interact with genotype and parity only in the executive functions memory composite as there was a large effect of age of menopause in carriers of the APOE ε4 allele compared to non-carriers.

Additionally, in individuals with four or more children an earlier age of menopause was associated with a greater reduction in executive functions composite scores, than in individuals with 0-3 children. Collectively, our findings suggest that earlier age of menopause is associated with impaired cognition, and E2-based MHT, depending on route of administration, is associated with improved cognitive performance, although this depends on cognitive domain. Transdermal E2-based MHT was associated with the largest positive effect on episodic memory, whereas oral E2 was associated with improved prospective memory. These results help clear up a number of inconsistencies in the literature on the influence of MHT on cognition in postmenopausal people.

### E2 MHT is associated with cognitive improvements based on cognitive domain tested

Our findings indicate that although increased menopause age and E2 MHT use positively influence episodic and prospective memory composite scores (functions associated with the medial temporal lobe including the hippocampus; (Afthinos et al., 2022), the effects on executive functions were less pronounced. This differential effect is not entirely surprising, especially given the distinct functions and ER distributions in these brain regions in both rats and mice (Mitra et al., 2003; Österlund et al., 1998). Previous work shows that ER density is increased post-menopause compared to pre-menopause in the middle frontal regions, posterior cingulate, pituitary, caudate and to a lesser extent, the inferior frontal regions (Mosconi et al., 2024). Contrastingly, the hippocampus did not show changes in ER density compared to pre menopause (Mosconi et al., 2024). This finding is interesting in light of the fact that we saw a larger positive effect size of E2 MHT in the episodic memory composite, which relies in part on the integrity of the hippocampus. However, ER density in the posterior cingulate and hippocampus was negatively associated with logical memory and trail making in postmenopausal individuals, suggesting that increased sensitivity or response to estrogens may be associated with impaired cognition in the frontal lobe (Mosconi et al., 2024). Therefore, this may explain why our work shows that E2 has a more robust effect on hippocampus-dependent tasks compared to those mediated by the frontal lobe. Indeed, other studies show that MHT (type not specified) was associated with greater brain activation in the left hippocampus and better verbal memory scores (Weber et al., 2013). These findings underscore the importance of considering specific cognitive domains and specific brain regions which may be differentially sensitive to E2 manipulations in postmenopausal people.

### E2 MHT is associated with cognitive improvements based on route of administration

Our novel finding that the positive associations on cognitive with E2-based MHT depends on the cognitive domain and route of administration, provides some clarity on the inconsistencies in the literature. Transdermal E2 was associated with improved scores on the episodic memory composite whereas oral E2 was associated with improved performance on the prospective memory composite compared to those who have never taken any MHT. Notably, no matter the route of administration, E2 was not associated with impaired performance on any of the cognitive domains tested compared to those that never took MHT. This is consistent with previous work showing that E2-based MHT, has either positive effects or neutral effects on cognition, brain health, and disease risk (Fischer et al., 2014; Kim et al., 2021; Maki & Resnick, 2000; Maki & Sundermann, 2009; Ryan et al., 2008, 2009; Wharton et al., 2011).

The route of E2 administration affects its metabolism, and therefore subsequently its bioavailability and dose, which will alter its efficacy. Our findings that transdermal E2 were associated with higher episodic memory scores could be because the transdermal administration of E2 bypasses first-pass hepatic metabolism, leading to higher and more stable plasma E2 levels, and potentially greater efficacy (Kuhl, 2005; Sitruk-Ware et al., 2007). However, transdermal E2 did not show a significant association with performance of prospective memory nor executive functions, which may be due to dose, type and availability of ERs in corresponding brain regions (see below). This latter finding is also in alignment with the findings of the KEEPS trial, which showed no significant effect of transdermal E2 on mostly executive functions, and on functions related to integrity of the frontal lobe (Gleason et al., 2015; Kantarci et al., 2018). Intriguingly, the KEEPS trial did find taking transdermal E2 was related to less steep declines than oral CEE in verbal memory, which relies on the integrity of the medial temporal lobe, although caution is warranted as these findings were not statistically significant (Gleason et al., 2015). Furthermore, the transdermal E2-treated group in their cohort had more white matter hyperintensities and a greater percentage APOE ε4 carriers than the placebo group at baseline, which may have masked the beneficial effects of transdermal E2 (Kantarci et al., 2018). Furthermore, it is possible our results differ from the KEEPS study as they did not control for the age of menopause in their analysis. Our data highlights the importance of considering the route of MHT administration, age of menopause, and investigation on the associations with different cognitive domains, which are often overlooked in investigations of MHT on the brain and cognition and doing so may account for inconsistencies in the literature.

That the method of administration of E2 has cognitive-domain specific effects could be explained by the differences in first-pass hepatic metabolism and resulting concentrations of systemic E2. In the CLSA data, oral E2 had a significant positive association on prospective memory compared to transdermal E2, an opposite pattern than seen with episodic memory. E2 has dose-dependent effects (Barha et al., 2009), and it is possible that a lower dose of E2 is required to show beneficial associations on prospective memory. Indeed, there is evidence that lower concentrations of E2 are associated with improved working memory or increased activation in the precentral gyrus which depends on dopamine availability (Dumas et al., 2018; Jacobs & D’Esposito, 2011). E2 infusions into the frontal cortex dose-dependently affect spatial working memory performance (Sinopoli et al., 2006). In addition, evidence from the animal literature suggests that E2 and other reproductive factors have stronger effects on neuroplasticity and inflammation in the hippocampus than in the frontal lobe (Eid et al., 2020; Lee, Cevizci, et al., 2024), indicating that doses of E2 have differential effects on frontal versus hippocampal mediated functions.

Our findings may also explain why animal studies more consistently find positive effects of E2 on cognition, whereas the evidence from human literature of MHT’s effects on cognition are more mixed. Work in animal models mostly investigates the effects of subcutaneous E2 (which is absorbed, metabolized, and dosed similarly to transdermal E2) on hippocampus dependent tasks perhaps more so than frontal-lobe based ones (L. A. Galea et al., 2001; J. Hao et al., 2007; Holmes et al., 2002; McClure et al., 2013; Talboom et al., 2008). Although subcutaneous and local infusions of E2 do positively influence memory performance of frontal-dependent tasks, they do so at different subcutaneous doses of E2 (Sinopoli et al., 2006; Wide et al., 2004). In the human literature, the majority of studies have examined oral doses of MHT on working memory or executive functions that largely depend on frontal-lobe integrity (Georgakis et al., 2016; Hogervorst et al., 2022; Kuh et al., 2018). It is important to acknowledge that dose-dependent effects may contribute to the observed outcomes in our present study, with a relatively lower dose (such as that delivered by oral E2) potentially offering greater benefits for frontal-lobe-dependent cognition, consistent with the animal literature (Wide et al., 2004).

In all our analyses, we included age at menopause as a covariate, recognizing its critical influence on cognition. Neglecting to account for this variable would confound the analysis of MHT effects, as differences in cognitive outcomes could be partially attributed to variations in the timing of menopause rather than the therapy itself. Therefore, adjusting for age at menopause for MHT’s effects on cognition, minimizing potential biases.

### Age of Menopause is associated with episodic and prospective memory performance

Our work, in line with some previous work, suggests that an earlier age of menopause is associated with worse cognitive performance across cognitive domains (Georgakis et al., 2016; Hogervorst et al., 2022; Kuh et al., 2018). Another study using the UK biobank found significant associations with menopause age on certain cognitive tasks (numerical recall, fluid intelligence and pairs matching) but not on prospective memory, although the means leaned in the same direction (Costantino et al., 2023). An earlier age of menopause being associated with reduced episodic and prospective memory performance suggests that prolonged exposure to ovarian hormones can confer neuroprotective benefits, on both the limbic system and parietal system. Taken together, these findings highlight the importance of considering both the age of menopause and its domain-specific effects on cognitive performance, offering valuable insights into the nuanced ways in which menopause timing may influence brain health and aging.

### Age of menopause influences on executive functions in APOEe4 carriers and those with grand parity

Further analyses explored the potential moderating effects of carrying the APOE ε4 allele and previous parity (pregnancy and motherhood) on cognitive performance. Our data showed that carrying the APOE ε4 allele and the grand parity (4 or more biological children) was associated with poorer executive functions composite scores with earlier age of menopause relative to their counterparts APOE ε4 carriers show altered lipid metabolism and increased vulnerability to AD with earlier loss of ovarian hormones (Valencia-Olvera et al., 2023). This may render carriers more sensitive to hormonal fluctuations, and therefore lead to our findings that greater benefits due to a later age of menopause compared to non-carriers. However, we find that carrying the APOE ε4 allele or the number of biological children did not modify the effects of E2-based MHT on episodic memory, prospective memory, or executive functions composite scores. Previous work on whether MHT’s effects depend on APOE status is equivocal, possibly due to lack of specificity in the type of MHT, and cognitive domain considered (reviewed in (Valencia-Olvera et al., 2023)).

Those with grand parity were more likely to show higher scores on executive functions with an older age of menopause than those with fewer or no children. This could be because multiple pregnancies result in prolonged exposure to high levels of endogenous placental hormones, which may have cumulative neuroprotective effects on the frontal lobe (Azcoitia et al., 2019; Saravia et al., 2007). It is also possible that this is because having four or more children may be associated with sleep debt, endocrine and immune changes that leave an individual especially vulnerable to the effects of earlier reductions in estradiol levels, but why this would only affect cognitive domains impacting the frontal lobe is not clear. ERs and PRs are distributed widely across the brain, and there is a menopause-associated increase in ERs in various regions, including frontal regions and caudate but intriguingly not in the hippocampus (Barkhoudarian & Kelly, 2017; Grahn et al., 2008; Mosconi et al., 2024).Higher parity could enhance neural resilience or promote neural plasticity in brain regions associated with executive functions resulting in greater benefits. Our present work indicates that the positive associations of exogenous E2 on episodic and prospective memory were independent of APOE status and parity.

Because both grand parity and APOE4 genotype are associated with greater risk for AD (Bae et al., 2020; Jung et al., 2020; Riedel et al., 2016), our results suggest there is a cognitive protection for both those at higher risk for AD prior to cognitive impairments. Although parity and APOE4 status did not influence the cognitive effects of E2-based MHT in the present study, they are still important considerations for effective, individualized, precision medicine, and may matter for other types of MHT (e.g., E1-based MHT).

### Limitations

Some study limitations include the following: For example, the sample of individuals from the CLSA taking MHTs was wealthier and predominantly white, more so than the Canadian population, limiting generalisability (Government of Canada, 2022). Additionally, we focused exclusively on E2-based MHT, which limits the applicability of our results to other MHT. Indeed, the effects of estrone-based MHT on cognition are reported to be largely detrimental or neutral (Andy et al., 2024; Coker et al., 2010; Espeland, 2004; Ryan et al., 2008). We also did not differentiate between individuals taking only E2, or E2 along with progesterone or progestins, as in individuals with an intact uterus, concurrent progesterone or progestins are needed to prevent endometrial hyperplasia or cancer (Griksiene et al., 2022; Sherwin & Grigorova, 2011). However, the proportion of individuals taking concurrent progesterone or progestins was matched between the transdermal E2 group and oral E2 group.

Furthermore, we carried out a sensitivity analysis for individuals taking progesterone or progestins, and determined that our effects remained robust. Additionally, we did not exclude females with hysterectomy in our analysis, as we did not have information on whether those who underwent the procedure did so before or after natural menopause. Hysterectomy is related to an earlier menopause onset by approximately 2-3 years, and an increased risk for early onset dementia (Phung et al., 2010). However, the proportion of females with hysterectomy were not statically different between the transdermal and oral E2 groups. Moreover, the proportion of females with hysterectomy was significantly higher in both MHT groups compared to the group not taking any MHT. If anything, hysterectomy should have worsened cognition in these groups, but this was not the case. In order to account for any confounding effects, we ran a sensitivity analysis by excluding those with hysterectomy, and our results remained robust. In this study, in order to control for menopause age in our model, when considering MHT use, we only examined postmenopausal individuals. In the future it would be interesting to examine those that are perimenopausal and taking MHT. Unfortunately, we were unable to consider gender diversity in this cohort, as gender was not queried in the baseline questionnaire.

Indeed, sex was only self-reported, and we included those who reported “female” sex at birth. It is important for future studies to examine how gender-diverse individuals may be affected by MHT. We were limited with the sample that we had available from the CLSA, but further research and study design should aim to include a broader range of ethnicities and more gender diversity when possible. Additionally, BMI was used as a covariate in our analyses as it was statically different across MHT groups but some have suggested this is an imperfect measure (Khanna et al., 2022). We are also unable to examine the influence of E2 dose in this cohort which would be an important future study. Finally, as our study is cross-sectional and observational, this could lead to selection bias in MHT use and residual confounding. As such, we cannot establish causality between menopause age, MHT use, and cognitive outcomes, and further longitudinal and interventional studies may be useful to confirm the associations we find and to elucidate any underlying mechanisms.

Despite these limitations, our study provides important insight into the effects of menopause timing and E2-based MHT on cognition, including the importance of considering both genetic factors and reproductive history when considering cognitive performance and aging in healthy females.

## CONCLUSIONS

Our findings show that the effects of E2-based MHT on cognition differ based on the route of administration and cognitive domain. Transdermal E2 was significantly associated with episodic memory, whereas oral E2 was associated with prospective memory scores. E2-based MHT was not associated with executive functions composite scores. An earlier age of menopause was associated with reduced cognitive performance in episodic and prospective memory domains. However, for executive functions, an earlier age of menopause was only associated with lower scores with grand parity (four or more children) and was more pronounced in APOE ε4 allele carriers. These results emphasize the importance of considering both cognitive domain and route of administration of E2 when evaluating the effects of MHT on cognition. Our study population consisted of only cognitively healthy individuals without diagnosed AD or depression, and our results align with the ‘healthy cell bias’, which suggests that the beneficial effects of hormones are most apparent in individuals with healthy neural systems (Brinton, 2005, 2008). This suggests that E2-based MHTs may be most effective as prophylactic measures rather than treatments for existing cognitive deficits. Our study provides clarity on inconsistencies in the literature, highlighting the influence of route of administration of MHT, and age of menopause can significantly affect cognitive outcomes dependent on cognitive domain. An improved understanding of how MHT factors influence cognitive health is crucial for developing targeted interventions to mitigate long-term neurological risks in females.

## Data Availability

Data are available from the Canadian Longitudinal Study on Aging (www.clsa-elcv.ca) for researchers who meet the criteria for access to de-identified CLSA data.

## Acknowledgements

This research was made possible using the data/biospecimens collected by the Canadian Longitudinal Study on Aging (CLSA). Funding for the Canadian Longitudinal Study on Aging (CLSA) is provided by the Government of Canada through the Canadian Institutes of Health Research (CIHR) under grant reference: LSA 94473 and the Canada Foundation for Innovation, as well as the following provinces, Newfoundland, Nova Scotia, Quebec, Ontario, Manitoba, Alberta, and British Columbia. This research has been conducted using the CLSA Comprehensive Baseline Dataset version 6.0, and Genome-wide Genetic Data – Version 3.0 under Application Number 20CA019. The CLSA is led by Drs. Parminder Raina, Christina Wolfson and Susan Kirkland. The opinions expressed in this manuscript are the author’s own and do not reflect the views of the CLSA.

We would like to thank Dr. Ulrike Mayer, and Dr. Arianne Albert for their help sorting and cleaning data for this manuscript.

---

**Table 1:** Demographics for Individuals using Oral E2, Transdermal E2, or who have never taken MHT.

